# Implementing genomic screening in diverse populations

**DOI:** 10.1101/2021.01.04.21249213

**Authors:** Noura S. Abul-Husn, Emily R. Soper, Giovanna T. Braganza, Jessica E. Rodriguez, Natasha Zeid, Sinead Cullina, Dean Bobo, Arden Moscati, Amanda Merkelson, Ruth J.F. Loos, Judy H. Cho, Gillian M. Belbin, Sabrina A. Suckiel, Eimear E. Kenny

## Abstract

**Background:** Population-based genomic screening has the predicted ability to reduce morbidity and mortality associated with medically actionable conditions. However, much research is needed to develop standards for genomic screening, and to understand the perspectives of people offered this new testing modality. This is particularly true for non-European ancestry populations who are vastly underrepresented in genomic medicine research. Therefore, we implemented a pilot genomic screening program in the Bio*Me* Biobank in New York City, where the majority of participants are of non-European ancestry.

**Methods:** We initiated genomic screening for well-established genes associated with hereditary breast and ovarian cancer syndrome (HBOC), Lynch syndrome (LS), and familial hypercholesterolemia (FH). We evaluated and included an additional gene (*TTR*) associated with hereditary transthyretin amyloidosis (hATTR), which has a common founder variant in African ancestry populations. We evaluated the characteristics of 74 participants who received results associated with these conditions. We also assessed the preferences of 7,461 newly enrolled Bio*Me* participants to receive genomic results.

**Results:** In the pilot genomic screening program, 74 consented participants received results related to HBOC (N=26), LS (N=6), FH (N=8), and hATTR (N=34). Thirty-three of 34 (97.1%) participants who received a result related to hATTR were self-reported African/African American (AA) or Hispanic/Latinx (HL), compared to 14 of 40 (35.0%) participants who received a result related to HBOC, LS, or FH. Among 7,461 participants enrolled after the Bio*Me* protocol modification to allow the return of genomic results, 93.4% indicated that they would want to receive results. Younger participants, women, and HL participants were more likely to opt to receive results.

**Conclusions:** The addition of *TTR* to a pilot genomic screening program meant that we returned results to a higher proportion of AA and HL participants, in comparison with genes traditionally included in genomic screening programs in the U.S. We found that the majority of participants in a multi-ethnic biobank are interested in receiving genomic results for medically actionable conditions. These findings increase knowledge about the perspectives of diverse research participants on receiving genomic results, and inform the broader implementation of genomic medicine in underrepresented patient populations.

## INTRODUCTION

The rapidly decreasing cost of genomic technology has accelerated the implementation of new genomic applications in medicine, including population-based genomic screening (1, 2). The rationale for genomic screening is that it can identify individuals at increased risk for serious health conditions that may otherwise be missed, and for which there are effective interventions to mitigate risk. Three conditions are designated by the Centers for Disease Control and Prevention (CDC) as Tier 1 genomic applications having the most evidence to support their early detection and intervention: hereditary breast and ovarian cancer (HBOC), Lynch syndrome (LS), and familial hypercholesterolemia (FH) (3). An estimated 1-2% of the population harbor a pathogenic or likely pathogenic (P/LP) variant in one of the nine genes underlying these conditions (2, 4-7). However, because at present these conditions are poorly ascertained in routine patient care, most affected individuals are not aware they are at risk (1, 2, 4, 5, 7).

There are many considerations and challenges to incorporating genomic screening into clinical practice, including clinical utility, patient and provider preferences, and healthcare infrastructure and resources to provide downstream care. Biobanks embedded in health systems offer excellent research environments to implement and assess genomic screening programs. In the next five years, genomic data for over 60 million patients is expected to be generated in biobanks in health systems globally (8). Some of these biobanks are built with or have pivoted toward a regulatory structure enabling the return of results to participants (9). However, most biobank programs that return genomic results are doing so in health systems serving predominantly European (EA)-ancestry patients (1, 10). Lack of diversity impedes research on how to best tailor genomic screening to multi-ethnic populations, and misses opportunities to learn about diverse patients’ preferences for receiving genomic results. As large-scale genomic and precision medicine research efforts, such as the All of Us Research Program (11), prioritize engagement of diverse populations, it is increasingly important to evaluate approaches to returning individual genomic results to these participants.

Here we describe a pilot genomic screening program to return results to consented participants of the Bio*Me* Biobank, an ongoing biorepository in New York City (NYC). Bio*Me* participants represent broad ancestral diversity, with over 65% self-reporting as non-EA (9), and with evidence of population-specific disease burdens (12). As the suitability of genomic results to be returned to biobank participants is the subject of ongoing deliberation (13), we followed an approach recently proposed by an expert working group of the Genomics and Population Health Action Collaborative, which outlines a tier system for the inclusion of genes in genomics-based screening programs (14). Our pilot program includes the CDC Tier 1 genes: *BRCA1* and *BRCA2* (associated with HBOC); *MLH1, MSH2, MSH6*, and *PMS2* (associated with LS); and *LDLR, APOB*, and *PCSK9* (associated with FH). Given the unique, multi-ethnic composition of the Bio*Me* Biobank, we established a process to evaluate additional genes that could benefit research participants historically underrepresented in genomic screening efforts. Consequently, results have been returned for *TTR* variants associated with hereditary transthyretin amyloidosis (hATTR), a condition predominantly affecting African ancestry populations in the U.S (15), with newly-approved therapeutics to improve health outcomes (16). We discuss the implementation of a pilot genomic screening program tailored to diverse patient populations, and participant preferences for receiving genomic results in a multi-ethnic biobank. We report characteristics of the first 74 participants to receive results, and consider the impact of including a condition disproportionately affecting minority populations.

## METHODS

### Study Population

The Bio*Me* Biobank is an electronic health record (EHR)-linked biorepository that has been enrolling participants non-selectively from across the Mount Sinai Health System (MSHS) since 2007. To date, there are over 55,000 participants enrolled in Bio*Me* under Institutional Review Board (IRB)-approved study protocol and consent. Bio*Me* participants consent to provide DNA and plasma samples linked to their de-identified EHRs, and to be recontacted for future research. Exome sequence data is available for 30,223 Bio*Me* participants aged 18 years or older (enrolled from September 2007 to November 2016), through a collaboration with the Regeneron Genetics Center (5). Self-reported race/ethnicity of participants is derived from a multiple-choice survey administered on enrollment in Bio*Me*, as previously described (12). In October 2018, the Bio*Me* protocol and consent were modified to allow participants the option to receive clinically confirmed, medically actionable genomic results.

### Pre-Pilot Bio*Me* Return of Results Survey

Prior to the Bio*Me* protocol modification allowing the return of results, we assessed adult participants’ perspectives on the return of genomic results through a 21-item survey study. The survey study was determined exempt by the Icahn School of Medicine at Mount Sinai’s IRB. Survey data were collected between June 2015 and February 2016. A link to the online survey was included in one edition of a newsletter mailed to Bio*Me* participants, and 500 randomly selected Bio*Me* participants were mailed a paper version of the survey and a pre-paid return envelope. Survey responses were anonymous. The survey assessed demographic and socioeconomic factors, participants’ interest in receiving genomic results, perceived benefits and concerns, and preferred method for result disclosure. Survey measures were guided by previous studies on the willingness of minority populations to participate in genomic research, genomic information needs among diverse biobank participants, and attitudes of health professionals and the public toward the return of incidental results (**Additional File 1: Table S1**) (17-20). Not all survey data are presented in this paper.

### Pilot Genomic Screening Program

A pilot genomic screening program was launched in February 2019 to disclose clinically confirmed, medically actionable genomic results to Bio*Me* participants. The program initially included HBOC, LS, and FH, and was expanded in November 2019 to include hATTR. Because participants with available exome sequence data were consented prior to the Bio*Me* protocol modification allowing the return of results, this limited the number of participants eligible for the pilot genomic screening program. Outreach to Bio*Me* participants to encourage updating of consent forms included unselected outreach via Bio*Me* Biobank recruitment events and newsletter announcements, as well as targeted outreach via mailed letters letting participants know about changes to the Bio*Me* protocol. At the time of this study, 692 participants aged 18 years or older with available exome sequence data had updated their Bio*Me* consents to indicate that they wished to receive results.

### Clinical Confirmation of Research Results

Exome sequence data from 692 eligible participants were screened for research results. Research results included P/LP variants in all 10 genes of interest identified by positional intersection with the ClinVar database (accessed August 2019) (21), and additional predicted loss-of-function (stop gain, start loss, frameshift, splice acceptor, or splice donor) variants in 7 genes for which loss of function is a known disease mechanism (i.e., *BRCA1, BRCA2, MLH1, MSH2, MSH6, PMS2*, and *LDLR*). In addition, we screened available genotype array data from eligible participants for V142I, a common pathogenic variant in *TTR*. Consented participants with a research result provided a new blood sample for clinical confirmation of the suspected finding. Research results were confirmed by Sanger sequencing at Sema4, a New York State-approved and CLIA-certified clinical genetic testing laboratory (22). The clinical laboratory generated individual reports that were reviewed by the study team. Only variants confirmed by Sanger sequencing and classified as P/LP for the four conditions included in the program were disclosed to participants.

### Result Disclosure

Consenting participants with clinically confirmed P/LP variants were scheduled for an in-person genetic counseling visit for result disclosure. These visits were transitioned to telemedicine in April 2020 due to the COVID-19 pandemic, which included video visits where possible and telephone encounters if technological barriers were insurmountable. Participants with prior knowledge of their genomic risk were informed of their result by telephone, and were offered an in-person genetic counseling visit. Genetic counseling visits were conducted with a third-party Spanish interpreter for Spanish-speaking participants. The components and content of result disclosure and genetic counseling were consistent across all service delivery models (see **Additional File 1: Figure S1**). Participants received a copy of their clinical report and an internally developed fact sheet (in English or Spanish) on the associated condition that summarized information received. After the visit, results were uploaded to the participant’s EHR and their problem list (list of patient diagnoses) was updated to include their gene variant status. All participants with telemedicine encounters received a follow-up phone call one week post-result disclosure.

### Bio*Me* Participants’ Preferences to Receive Results

We evaluated Bio*Me* participants’ consent data during the first year following the change in Bio*Me* protocol allowing participants the option to receive results (October 2018 - October 2019). We queried 7,461 newly enrolled participants’ responses to the question “Do you wish to receive genetic results?” We analyzed differences in responses by sex, age, and self-reported race/ethnicity categories, excluding Native Americans (N = 9) and those with missing race/ethnicity information (N = 268).

### Data Analysis

Data were analyzed using JMP Version 15 (SAS Institute Inc). Counts and percentages were calculated for categorical variables, and medians and ranges were calculated for continuous variables. Pearson’s chi-squared test and multiple logistic regression were performed to test for statistical differences across groups. *P* < 0.05 was considered statistically significant.

## RESULTS

### Pre-Pilot Survey to Understand Bio*Me* Participants’ Preferences to Receive Genomic Results

Seventy-two Bio*Me* participants responded to a survey assessing perspectives on the return of hypothetical genomic results (34 by mail and 38 electronically). Compared to all Bio*Me* participants enrolled at the time, survey respondents were younger and more likely to be born in the U.S. Respondents were 58.3% non-EA by self-report, which is broadly representative of the diversity of Bio*Me* participants (**Additional File 1: Table S2)**. The majority replied that they definitely (73.9%) or probably (15.9%) still would have enrolled into Bio*Me* if genomic results were returned as part of the program, and the rest (10.1%) were unsure (**Additional File 1: Figure S2A**). Participants were asked to select among reasons to receive or not receive results (**Additional File 1: Figure S2B**). Reasons to receive results included to help themselves (87.1%), help their family (75.7%), help with family planning (40.0%), and feeling ownership over results (34.3%). Reasons to not receive results included discrimination concerns (54.3%), anxiety (45.7%), privacy concerns (28.6%), and the inability to make health changes (21.4%). The majority (54.5%) preferred to receive all genomic results, and 31.8% preferred results about specific diseases that genetics experts think are important (**Additional File 1: Figure S2C**). Only 3.0% indicated that they would not want to receive any type of result. Most preferred to receive results from a genetic counselor (45.7%; **Figure 1A**). Preferred modes of result disclosure were by a letter in the mail (50.0%) or in person (38.6%; **Figure 1B**). Results from this survey helped inform the change in the Bio*Me* protocol to allow participants the option to receive genomic results and the implementation of a pilot genomic screening program.

**Figure 1.**
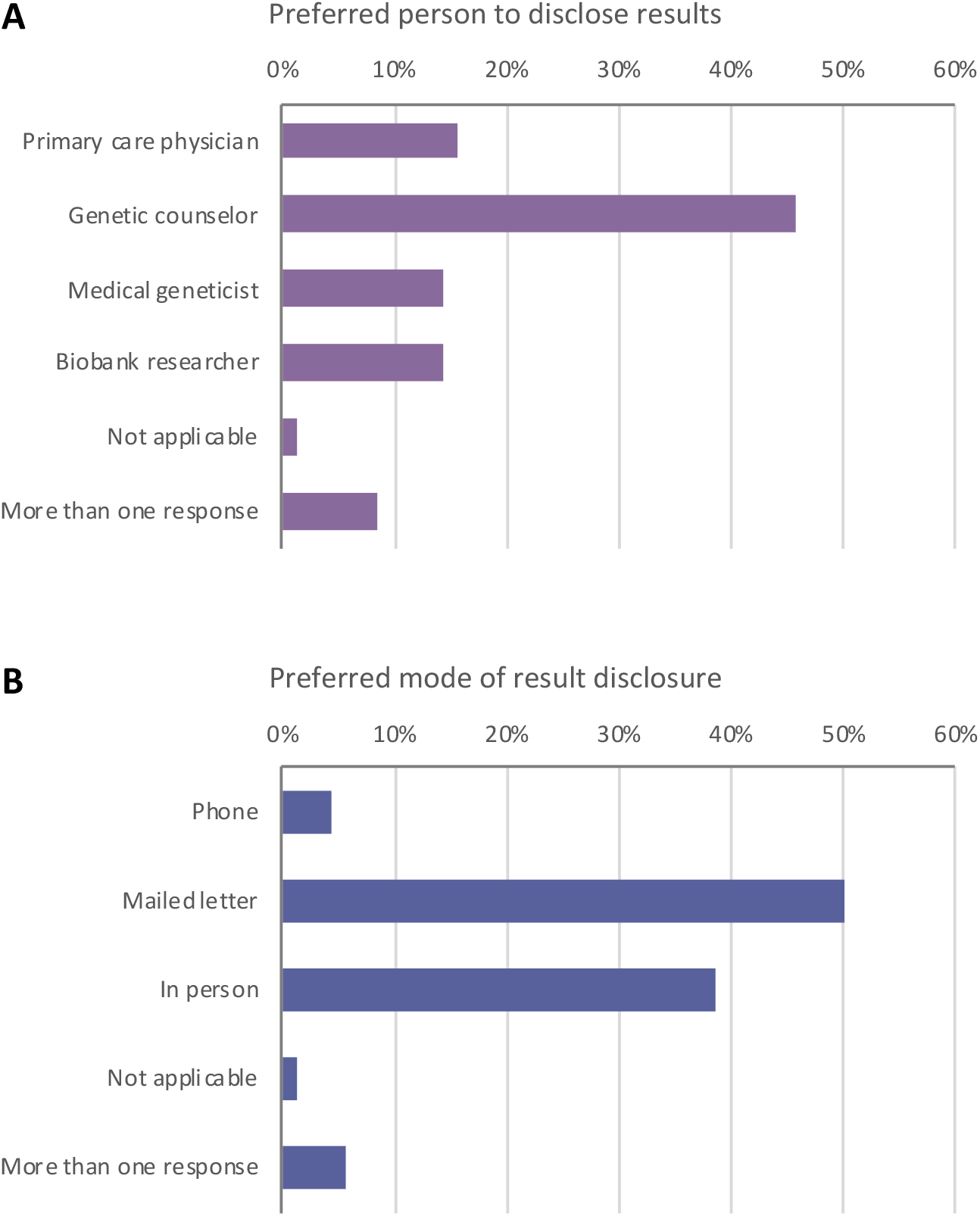
Previously enrolled Bio*Me* participants’ (N=72) survey responses regarding (A) preferred person to disclose genomic results and (B) preferred mode of results delivery. Participants were most likely to prefer that a genetic counselor (45.7%) disclose results, and that results be returned by mailed letter (50.0%) or in person (38.6%).

### Process for Inclusion of Genes and Conditions in a Pilot Genomic Screening Program

The pilot genomic screening program initially included nine genes associated with HBOC, LS, and FH (3). A GenomicsFirst Committee (comprising medical geneticists, genetic counselors, and non-genetics specialists with expertise in hereditary conditions) was concurrently established to evaluate additional genes for result return. We developed a process to evaluate genes based on 1) prevalence of P/LP variants in Bio*Me*, 2) current evidence for medical actionability, and 3) availability and engagement of appropriate specialty care at MSHS. The GenomicsFirst Committee recommended the addition of hATTR based on the high prevalence and clinical impact of *TTR* V142I in Bio*Me* (15), review of the Clinical Genome Resource (ClinGen) Actionability Adult Summary Report (23, 24), availability of new non-invasive procedures for the diagnosis of cardiac amyloidosis (25), FDA approval of new therapies for cardiac and neuropathic transthyretin amyloidosis (16, 26, 27), and the availability of expert multidisciplinary clinical care for amyloidosis at MSHS.

### Implementation of a Pilot Genomic Screening Program

Adult Bio*Me* participants with available exome sequence data who had updated their consent and indicated that they wished to receive results (N = 692) were eligible for inclusion in the pilot genomic screening program (**Figure 2**). Ninety-four participants had a research result, one of whom opted out of receiving results. Among the 93 remaining participants, results from 78 were confirmed by Sanger sequencing and were classified as P/LP by the clinical genetic testing laboratory. The others were not confirmed by Sanger sequencing (N = 3), or were downgraded (N = 11 classified as variants of uncertain significance, and N = 1 *APOB* variant associated with hypobetalipoproteinemia). All Sanger confirmed P/LP and downgraded variants are listed in **Additional File 1: Table S3**.

**Figure 2.**
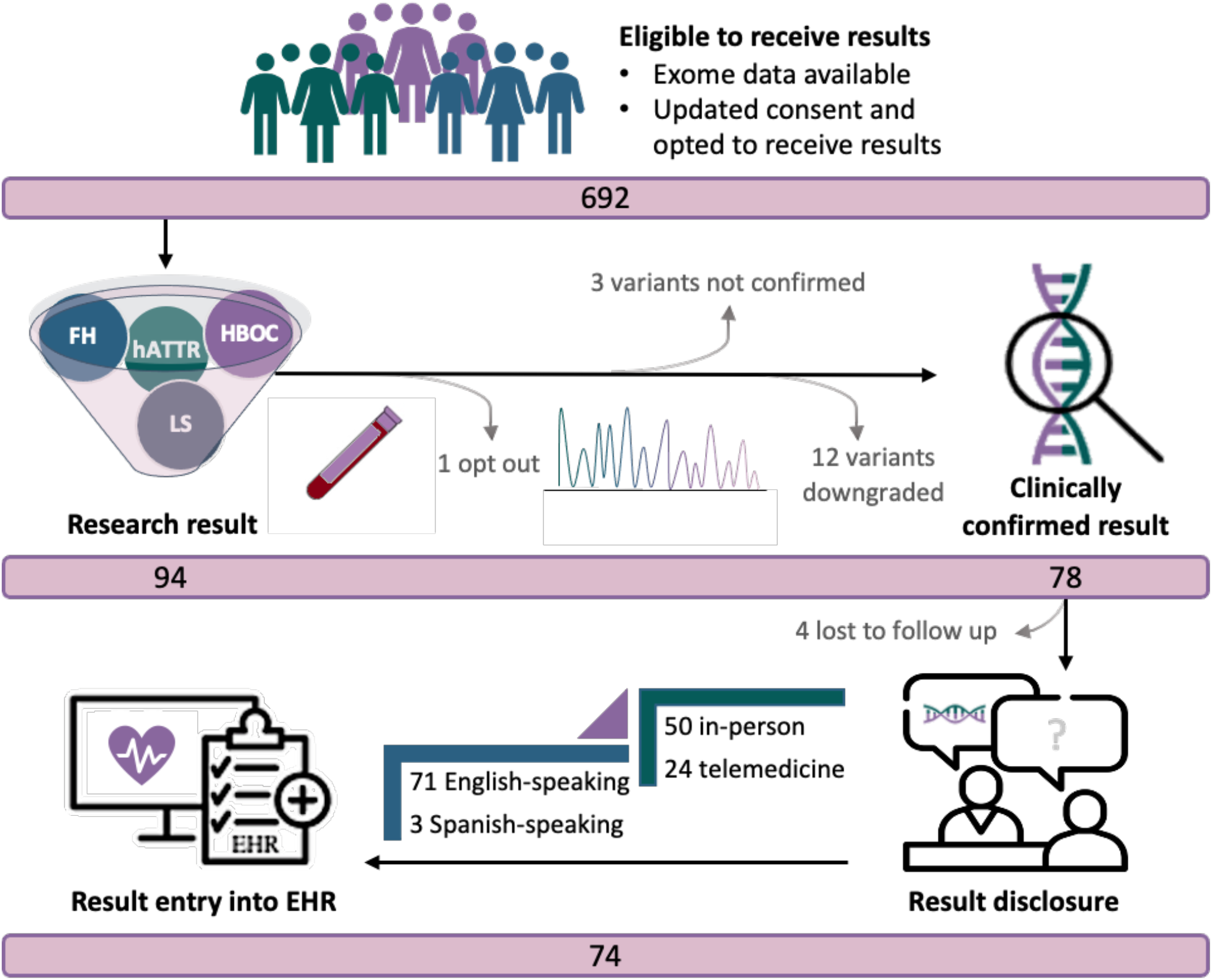
Pilot genomic screening program. Schematic outlining a pilot genomic screening program to return clinically confirmed pathogenic and likely pathogenic (P/LP) variants in genes related to hereditary breast and ovarian cancer syndrome (HBOC), Lynch syndrome (LS), familial hypercholesterolemia (FH), and hereditary transthyretin amyloidosis (hATTR).

Seventy-four of 78 participants with clinically confirmed P/LP variants received results (**Figure 2**). Four participants (5.3%) did not respond to numerous outreach attempts and therefore have not received results to date. Participants who received results were predominantly women (82.4%), and their median age was 58 years (age range 28 – 83 years; **Table 1**). We compared self-reported race/ethnicity categories of participants receiving a genomic result associated with a CDC Tier 1 condition (HBOC, LS, and FH) *vs*. hATTR. The proportion of AA or HL individuals receiving results through this program increased from 35.0% to 63.5% with the addition of *TTR* (chi square p = 3.6×10^−3^).

**Table 1.** Characteristics of Bio*Me* participants eligible to receive results and receiving results through a pilot genomic screening program.

| Participant demographics and characteristics | Eligible to receive results<br>(N = 692) | Received P/LP results |  |  | Downgraded variants<br>(N = 12) |
| --- | --- | --- | --- | --- | --- |
|  |  | CDC tier 1 (N = 40) | hATTR (N = 34) | CDC tier 1 + hATTR (N = 74) |  |
| Age, median (range) | 60 (18 - 95) | 61 (28 - 83) | 57 (30 - 80) | 58 (28 - 83) | 61 (30 - 76) |
| Female sex, N (%) | 458 (66.2) | 33 (82.5) | 28 (82.4) | 61 (82.4) | 8 (66.7) |
| Self-reported ancestry, N (%) |  |  |  |  |  |
| African American/African | 156 (22.5) | 6 (15.0) | 18 (52.9) | 24 (32.4) | 3 (25.0) |
| East/Southeast Asian | 26 (3.8) | 2 (5.0) | 0 | 2 (2.7) | 3 (25.0) |
| European | 186 (26.9) | 20 (50.0) | 1 (2.9) | 21 (28.4) | 5 (41.7) |
| Hispanic/Latinx | 249 (36.0) | 8 (20.0) | 15 (44.1) | 23 (31.1) | 0 |
| Native American | 1 (0.1) | 0 | 0 | 0 | 0 |
| Other | 41 (5.9) | 3 (7.5) | 0 | 3 (4.1) | 1 (8.3) |
| South Asian | 9 (1.3) | 0 | 0 | 0 | 0 |
| Multiple selected | 22 (3.2) | 1 (2.5) | 0 | 1 (1.4) | 0 |
| Not available | 2 (0.3) | 0 | 0 | 0 | 0 |
| Spanish as preferred language, N (%) | - | 0 | 3 (8.8) | 3 (4.1) | - |
| Prior knowledge of genomic risk, N (%) | - | 14 (35.0) | 1 (2.9) | 15 (20.3) | - |
| Mode of result disclosure, N (%) | - |  |  |  | - |
| In-person |  | 24 (60.0) | 26 (76.5) | 50 (67.6) |  |
| Telemedicine |  | 16 (40.0) | 8 (23.5) | 24 (32.4) |  |

<sup>a</sup>Includes individuals with previous knowledge of their genomic result

The majority (79.7%) of participants who received results were not previously aware of their genomic risk. Among 15 participants with prior knowledge of their genomic risk, 13 had a result associated with HBOC and 2 with a non-HBOC condition (hATTR and FH). The proportion of participants receiving a genomic result that they were previously unaware of increased from 65.0% to 79.7% with the addition of *TTR* (chi square *p* = 0.08). Notably, the individual with a prior diagnosis of hATTR was the only EA individual to receive a *TTR* result, and the only individual not harboring V142I. Three of 34 (8.8%) *TTR* results were disclosed with a third-party Spanish interpreter to participants whose preferred language was Spanish.

### Newly Enrolled Bio*Me* Participants’ Preferences to Receive Genomic Results

To understand participants’ preferences to receive genomic results at a larger scale and inform the broader implementation of genomic screening in Bio*Me*, we evaluated the consents of newly enrolled Bio*Me* participants in the one-year period following protocol modification (**Figure 3**). Among 7,461 newly enrolled participants, 6,968 (93.4%) indicated that they wished to receive genomic results. Preference to receive results was greater in women than men (94.2% *vs*. 92.2%, chi square *p* = 6.8×10^−4^) and in younger participants (chi square *p* = 4.3×10^−13^). Preference varied by self-reported race/ethnicity (chi square *p* = 6.2×10^−11^), with a greater proportion of HL individuals (95.9%) opting to receive results compared to other groups. In a multivariate model including age, sex, and self-reported race/ethnicity, all three predictors remained significantly associated with return of result preference (*p* = 2.7×10^−20^).

**Figure 3.**
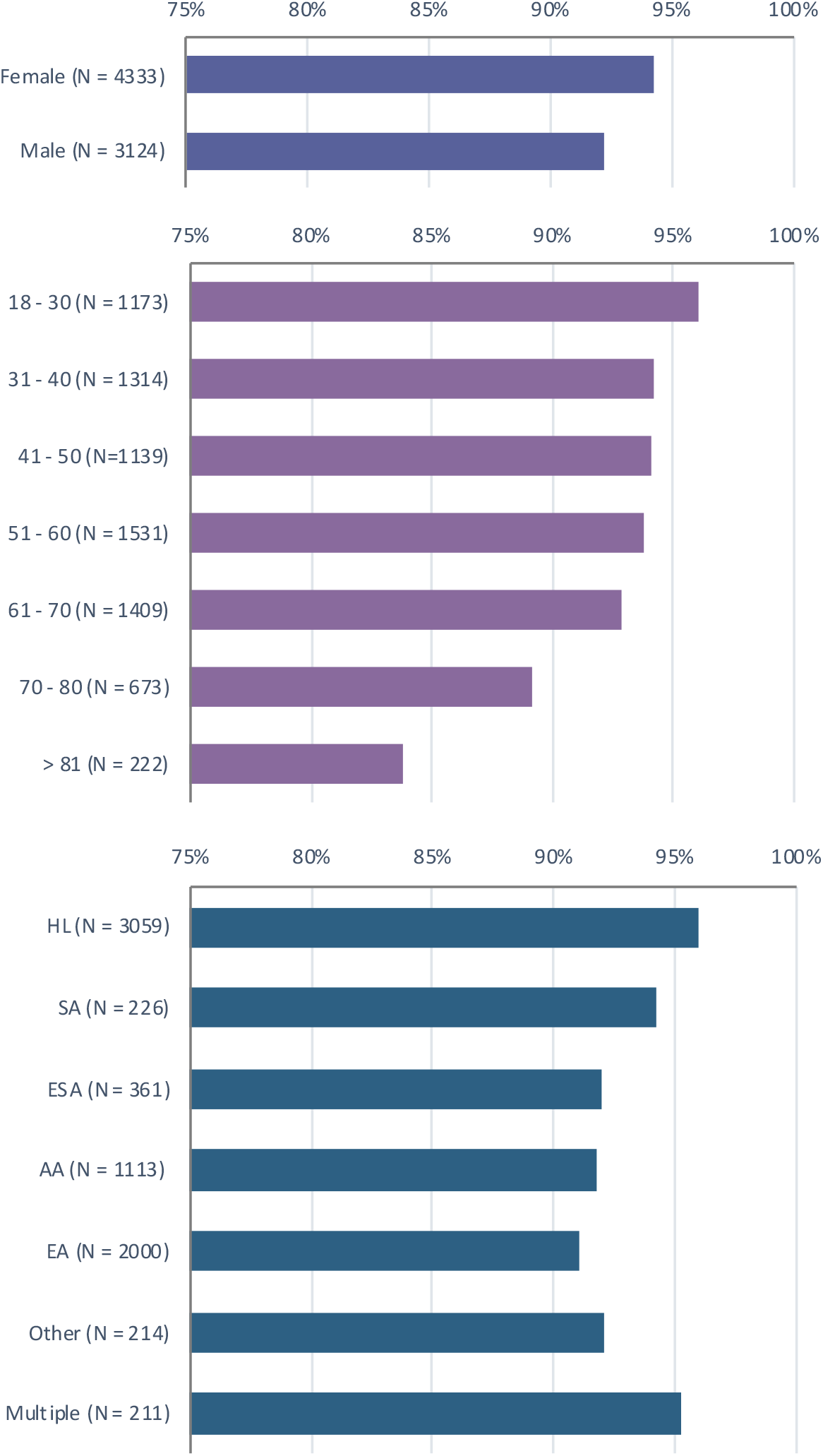
Preferences for receiving genetic results among 7,461 newly enrolled Bio*Me* participants across sex, age, and self-reported race/ethnicity groups. 94.2% of females vs. 92.2% of males opted to receive results (chi square p = 6.8×10^−4^). Younger participants were significantly more likely to prefer to receive results than older participants (chi square p = 4.3×10^−13^). HL individuals (95.9%) were more likely to opt to receive results, compared to other race/ethnicity groups (chi square p = 6.2×10^−11^). AA, African American; ESA, East/Southeast Asian; HL, Hispanic/Latinx; EA, European; SA, South Asian.

## DISCUSSION

Population genomic screening provides an opportunity to identify individuals at elevated risk of diseases for which preventive measures or treatments exist, and that may be underrecognized by standard clinical approaches (4, 5, 7, 15). This promotes proactive care by informing asymptomatic or presymptomatic individuals of their elevated genomic risk for disease and engaging them in personalized risk management (28, 29). We sought to improve our understanding of diverse biobank participants’ preferences with respect to receiving genomic results, and to implement a pilot genomic screening program tailored to better serve minority patient populations who are vastly underrepresented in genomic research and genomic medicine applications (30-32).

A pre-pilot survey of Bio*Me* participants suggested broad interest in receiving personal genomic results, and helped inform changes to the Bio*Me* protocol to allow participants this option. Survey respondents endorsed the return of all genomic results, including those that are uncertain or uninterpretable, which is similar to other studies reporting participants’ interest in receiving all personal genomic information (33). Consistent with most biobanks returning results, we elected to offer the return of medically actionable genomic results, which are most likely to positively impact clinical care because they are highly penetrant and are associated with conditions for which treatment and/or prophylactic measures are available (34, 35). The American College of Medical Genetics and Genomics (ACMG) lists 59 genes for reporting as incidental findings from clinical or research exome and genome sequencing studies based on medical actionability (35). However, the use of these genes for other purposes, including population screening, is not endorsed due to uncertain penetrance in asymptomatic individuals (36). Three conditions included in the ACMG list, HBOC, LS, and FH, are highly penetrant, poorly ascertained in clinical practice, and have established and effective interventions to prevent or mitigate disease risk. These were recently highlighted by an expert working group to meet Tier 1 guidelines for pilot genomic screening implementation (14). The same group suggested the inclusion of additional Tier 2 genes that are compelling for result return due to specific features of the study population, expertise within the study team, availability and quality of secondary screening, or other valid considerations. This approach, to start with the well-vetted Tier 1 conditions then prioritize additional genes and conditions specific to patient populations and health systems, is key to the responsible implementation of genomic screening programs in diverse settings.

In keeping with this approach, and aiming to address the paucity of genomic medicine applications in diverse, non-EA populations, we launched a pilot genomic screening program to return results related to CDC Tier 1 conditions and hATTR. To our knowledge, this is the first program to return *TTR* results to biobank participants. The decision to include *TTR* was multifactorial. A large number of Bio*Me* participants are heterozygous for *TTR* V142I (15), a common founder variant that is highly prevalent in AA and HL populations and significantly increases risk for cardiomyopathy and heart failure (37). With the availability of recently approved targeted therapies, hATTR is a medically actionable condition for which early diagnosis can guide treatment, thereby altering the disease course (16, 26, 27). It is an underrecognized and underdiagnosed condition (38), which supports the potential benefit of a genomics-first approach to identify individuals at risk for hATTR (39). The availability of multidisciplinary care for patients with or at risk for amyloidosis at MSHS further supported the inclusion of *TTR* in the program. The majority (97.1%) of participants who received a result related to hATTR were AA or HL, and Spanish was the preferred language of three of these participants. This compared to 35.0% of participants (none of whom were Spanish-speaking) who received a result related to HBOC, LS, or FH. Thus, the addition of hATTR to a genomic screening program in a diverse biobank introduced a higher proportion of AA, HL, and Spanish-speaking individuals in comparison with conditions traditionally included in return of results programs in the U.S.

Following the Bio*Me* protocol modification allowing participants the option to receive results, the vast majority of newly enrolled participants indicated that they wished to receive genomic results during the consent process, which was consistent with our pre-pilot survey. Participant preferences to receive results varied across sex, age and self-reported race/ethnicity groups. Consistent with previous studies, younger participants were more likely to prefer to receive results (40). A potential explanation for this is that the perceived personal utility of receiving genomic results to inform health risk management may decrease in older individuals. We also observed a higher proportion of HL participants consenting to receive genomic results compared to other self-reported race/ethnicity groups. In a small qualitative study of HL individuals in NYC, many participants cited the possibility of receiving actionable results as the primary reason to consider testing (41). However, participants also expressed concerns that by receiving actionable information, they may be forced to make unwanted or difficult lifestyle changes, which may be a barrier to the adoption of genomic screening. The views of HL populations have not been adequately explored in genomic research, potentially leading to suboptimal engagement of these individuals in genomic medicine. Our work helps to address this knowledge gap by providing insight into the preferences of HL populations for receiving genomic results.

There are limitations to this study. The first is the difficulty in outreaching to biobank participants who previously consented over the 13-year history of the biobank, in order to update their consents and indicate whether or not they would want to receive genomic results. This outreach is resource intensive, as participants are often not reachable due to outdated contact information and/or having left the health system; therefore, only a low volume of previously consented participants had updated their consent forms at the time of the pilot genomic screening program. A proportion of participants (20.3%) who received genomic results had previous knowledge of their personal genomic risk, particularly those at risk for HBOC (50.0%). In contrast, we previously found that only 26.6% of 218 Bio*Me* participants at risk for HBOC had EHR evidence of awareness of their genomic risk (5). This suggests that the population included in the pilot genomic screening program was enriched for participants with previous clinical genetic testing. A potential explanation for this is that individuals with previous genetic testing experience have greater appreciation of the benefits of receiving genomic results and were more inclined to update their Bio*Me* consents once the option to receive results was offered. Four out of 78 participants eligible to receive results were unable to be reached, suggesting barriers to re-establishing contact with some participants in order to disclose genomic results. To ensure adequate representation of hard-to-reach populations in genomic research, efforts to facilitate recontact are needed, such as ensuring multiple and preferred modes of communication, and maintaining contact and engagement with participants over time (42). The benefits of returning genomic research results, particularly in diverse populations and healthcare settings, is not well understood. Long-term follow up of participants is needed to determine the health-related and psychosocial impact of receiving results.

## CONCLUSIONS

This study details the implementation of a pilot genomic screening program in a highly diverse biobank in NYC. In keeping with pre-pilot survey findings endorsing a strong preference to receive genomic results, Bio*Me* participants overwhelmingly select this option regardless of age, sex, and self-reported race/ethnicity. The addition of *TTR* to our program increased the number of AA, HL, and Spanish-speaking participants receiving results, suggesting that including medically actionable genomic conditions with higher prevalence in non-EA populations can be an effective tool for expanding genomic medicine applications to historically underrepresented patient populations.

## Data Availability

Clinically confirmed pathogenic, likely pathogenic, and downgraded variants reported in this paper are tabulated in Additional File 1: Table S3. Exome sequencing and genotyping of Bio*Me* Biobank participants was performed in collaboration with the Regeneron Genetics Center. Individual-level data generated via this collaboration are not publicly available due to the terms of the Bio*Me* biospecimen and data access agreement, but may be requested directly from the corresponding author.

## DECLARATIONS

### Ethics Approval and Consent to Participate

The Bio*Me* Biobank is an ongoing research biorepository approved by the Icahn School of Medicine at Mount Sinai’s IRB (protocol number 07-0529). The Icahn School of Medicine at Mount Sinai’s IRB approved the pre-pilot survey study (protocol number 15-00440), including a waiver of informed consent and a HIPAA waiver of authorization. This research conformed to the Declaration of Helsinki.

### Availability of Data and Materials

Clinically confirmed pathogenic, likely pathogenic, and downgraded variants reported in this paper are tabulated in **Additional File 1: Table S3**. Exome sequencing and genotyping of Bio*Me* Biobank participants was performed in collaboration with the Regeneron Genetics Center. Individual-level data generated via this collaboration are not publicly available due to the terms of the Bio*Me* biospecimen and data access agreement, but may be requested directly from the corresponding author.

### Author Contributions

N.S.A.-H. and E.E.K. conceived and supervised all aspects of the study. N.S.A-H. and N.Z. designed the pre-pilot survey. N.S.A-H., E.R.S., A.M., S.A.S., and E.E.K. designed the pilot genomic screening program. G.M.B, S.C., A.M., and D.B. performed sequence data quality control and annotation. G.T.B., J.E.R., N.Z., and A.M. contributed to data acquisition. N.S.A-H., E.R.S., R.J.F.L., J.H.C., S.A.S., and E.E.K. contributed to data analysis and interpretation. N.S.A-H., E.R.S., G.T.B., S.A.S., and E.E.K. drafted the manuscript. All authors read and approved the final manuscript.

## Acknowledgements

We thank participants of the Bio*Me* Biobank for their permission to use their health and genomic information. We thank members of the GenomicsFirst Committee for their valuable input: Manisha Balwani, MD; George Diaz, MD, PhD; Amy Kontorovich, MD, PhD; Aimee Lucas, MD; Randi Zinberg, MS, CGC.

## ADDITIONAL FILES

**Additional File 1: Supplementary Figures and Tables** (PDF 274 kb)

**Table S1**. Questions from a return of genomic results preference survey administered to previously enrolled Bio*Me* Biobank participants.

**Table S2**. Demographic characteristics of 72 Bio*Me* participants who were respondents of a return of genomic results preference survey.

**Table S3**. Clinically confirmed pathogenic, likely pathogenic, and downgraded variants in a pilot genomic screening program.

**Fig. S1**. A model for returning genomic results to biobank research participants without prior knowledge of genomic risk.

**Fig. S2**. Pre-pilot survey to understand Bio*Me* participants’ (N = 72) preferences regarding the hypothetical return of results.

