## Supplementary Information for "Implementing genomic screening in diverse populations"

### ADDITIONAL FILE 1

[1]The Institute for Genomic Health, Icahn School of Medicine at Mount Sinai, New York, NY
[2]Department of Medicine, Icahn School of Medicine at Mount Sinai, New York, NY
[3]Department of Genetics and Genomic Sciences, Icahn School of Medicine at Mount Sinai, New York, NY
[4]GeneDx
[5]The Charles Bronfman Institute for Personalized Medicine, Icahn School of Medicine at Mount Sinai, New York, NY

**Corresponding Author:**
Noura S. Abul-Husn, M.D., Ph.D.
Icahn School of Medicine at Mount Sinai
One Gustave L. Levy Place, Box 1003
New York, NY 10029


**Supplementary Tables:**

1. **Table S1.** Questions from a return of genomic results preference survey administered to previously enrolled Bio*Me* Biobank participants.

2. **Table S2.** Demographic characteristics of Bio*Me* participants who were respondents of a return of genomic results preference survey (N = 72), and characteristics of all adult Bio*Me* participants enrolled at the time of the survey (N = 31,213).

3. **Table S3.** Clinically confirmed pathogenic, likely pathogenic, and downgraded variants in a pilot genomic screening program.

**Supplementary Figures:**

1. **Fig. S1. A model for returning genomic results to biobank research participants without prior knowledge of genomic risk.** After genomics-first identification of variant positive individuals, they receive genetic counseling and are then referred to specialty provider(s) for further evaluation and initiation of risk management.

2. **Fig. S2. Pre-pilot survey to understand Bio*Me* participants' (N = 72) preferences regarding the hypothetical return of results. A)** Decision to enroll into biobank research if the program returned genomic results. **B)** Selected reasons for receiving or not receiving genomic results. **C)** Selected categories of genomic results participants would want to receive.

**Table S1.** Questions from a return of genomic results preference survey administered to previously enrolled Bio*Me* Biobank participants.

| Question | Answer options |
|---|---|
| If, during your enrollment in *BioMe*, the recruiter told you that you **COULD** receive genetic research results from your participation, do you think you still would have enrolled? | • Definitely yes<br>• Probably yes<br>• Not sure<br>• Probably not<br>• Definitely not |
| What do you think are good reasons to receive genetic research results? (Check all that apply) | • To help myself (for example, by finding out what diseases I am at risk for)<br>• To help my family (for example, by finding out what diseases could run in the family)<br>• To help me make decisions about having children (for example, by finding out what diseases could be passed on to them)<br>• I feel ownership of my genetic research results<br>• Other (please specify) |
| What do you think are good reasons **NOT** to receive genetic research results? (Check all that apply) | • Concerns about my privacy<br>• Concerns about discrimination (health insurance, life insurance, employment, or other)<br>• Cannot make health changes based on genetic research results<br>• Receiving genetic research results would cause me or my family anxiety<br>• Other (please specify) |
| If given the choice, what type of genetic research results would you want to receive? | • Only genetic research results that the researcher thinks are important<br>• Only genetic research results about specific diseases that I think are important<br>• Only genetic research results about specific diseases that experts in genetics think are important and should be received by everyone<br>• All genetic research results, including those that are uncertain and/or cannot be interpreted at the moment<br>• None (I would not want to receive genetic research results) |
| If you were to receive genetic research results, how would you want to be informed? | • By phone<br>• By a letter in the mail<br>• In person<br>• Not applicable (I would not want to receive genetic research results) |
| If you were to receive genetic research results, who would you want to inform you? | • My primary care doctor that I see regularly for my healthcare<br>• A genetic counselor (a health professional trained in genetic diseases, particularly in returning and discussing implications of genetic testing results)<br>• A medical geneticist (a doctor who is specially trained in diagnosing and treating genetic diseases)<br>• A *BioMe* Biobank researcher or research coordinator<br>• Not applicable (I would not want to receive genetic research results) |

**Table S2.** Demographic characteristics of Bio*Me* participants who were respondents of a return of genomic results preference survey (N = 72), and characteristics of all adult Bio*Me* participants enrolled at the time of the survey (N = 31,213).

| Demographic characteristics (number of survey respondents) | Survey respondents N (%) | Bio*Me* participants N (%) | Chi-squared P value |
|---|---|---|---|
| Female (72) | 43 (59.7) | 18449 (59.1) | 0.9 |
| Age range (72) | | | < 0.0001 |
| 18 to 30 years | 27 (37.5) | 2149 (6.9) | |
| 31 to 50 years | 16 (22.2) | 8823 (28.3) | |
| 51 to 70 years | 26 (36.1) | 13538 (43.4) | |
| 71 years and over | 3 (4.2) | 6703 (21.5) | |
| Born in the U.S. (71) | 62 (87.3) | 19692 (63.1) | < 0.0001 |
| Self-reported ancestry (72) | | | 0.07 |
| African American/African | 9 (12.5) | 7670 (24.6) | |
| East/Southeast Asian | 2 (2.8) | 866 (2.8) | |
| Hispanic/Latinx | 24 (33.3) | 11072 (35.5) | |
| European | 30 (41.7) | 9459 (30.3) | |
| Native American, Other, or Multiple Selected | 7 (9.7) | 2146 (6.9) | |
| Jewish (70) | 20 (28.6) | - | - |
| Identify as religious (69) | 39 (56.5) | - | - |
| Have children (70) | 26 (37.1) | - | - |
| Highest level of school completed or highest degree (70) | | - | - |
| Less than high school degree | 1 (1.4) | | |
| High school degree or equivalent (e.g. GED) | 4 (5.7) | | |
| Some college but no degree | 16 (22.9) | | |
| Associate degree | 4 (5.7) | | |
| Bachelor degree | 26 (37.1) | | |
| Graduate degree | 19 (27.1) | | |
| Average household income (66) | | - | - |
| Less than $20,000 | 9 (13.6) | | |
| $20,000-$39,000 | 6 (9.1) | | |
| $40,000-$59,000 | 14 (21.2) | | |
| $60,000-$79,000 | 9 (13.6) | | |
| $80,000-$149,000 | 10 (15.1) | | |
| $150,000 or more | 18 (27.2) | | |

**Table S3.** Clinically confirmed pathogenic, likely pathogenic, and downgraded variants in a pilot genomic screening program.[a]

| Gene | CHR:POS:REF:ALT | cDNA Position | Protein Position | # Heterozygous Carriers | Interpretation |
|---|---|---|---|---|---|
| *BRCA1* | 17:43057062:T:TG | c.5266dupC | p.Gln1756fs | 2 | Pathogenic |
| | 17:43124027:ACT:A | c.68_69delAG | p.Glu23fs | 4 | Pathogenic |
| | 17:43094472:C:T | c.1059G>A | p.Trp353Ter | 1 | Pathogenic |
| | 17:43063917:A:C | c.5109T>G | p.Tyr1703Ter | 1 | Pathogenic |
| | 17:43049191:TG:T | c.5335delC | p.Gln1779fs | 1 | Pathogenic |
| | 17:43092615:TC:T | c.2915delG | p.Gly972fs | 1 | Pathogenic |
| | 17:43091455:T:TGC | c.4074_4075dupGC | p.Gln1359fs | 1 | Likely pathogenic |
| *BRCA2* | 13:32340128:C:T | c.5773C>T | p.Gln1925Ter | 1 | Pathogenic |
| | 13:32338277:G:T | c.3922G>T | p.Glu1308Ter | 2 | Pathogenic |
| | 13:32319298:G:T | c.289G>T | p.Glu97Ter | 1 | Pathogenic |
| | 13:32380085:C:T | c.9196C>T | p.Gln3066Ter | 1 | Pathogenic |
| | 13:32340300:GT:G | c.5946delT | p.Ser1982fs | 3 | Pathogenic |
| | 13:32336781:T:G | c.2426T>G | p.Leu809Ter | 1 | Pathogenic |
| | 13:32338783:CA:C | c.4429delA | p.Ile1477fs | 1 | Pathogenic |
| | 13:32370955:G:A | c.8488-1G>A | | 1 | Pathogenic |
| | 13:32339489:G:T | c.5134G>T | p.Gly1712Ter | 1 | Pathogenic |
| | 13:32340836:GACAA:G | c.6486_6489delACAA | p.Lys2162fs | 1 | Pathogenic |
| | 13:32336684:G:GA | c.2330dupA | p.Asp777fs | 1 | Pathogenic |
| | 13:32333282:G:T | c.1804G>T | p.Gly602Ter | 1 | Pathogenic |
| *MLH1* | 3:37012098:C:T | c.676C>T | p.Arg226Ter | 1 | Pathogenic |
| *MSH6* | 2:47799823:TC:T | c.1842delC | p.Cys615fs | 1 | Pathogenic |
| *PMS2* | 7:6005918:C:A | c.137G>T | p.Ser46Ile | 1 | Pathogenic |
| | 7:5986838:G:A | c.1927C>T | p.Gln643Ter | 1 | Pathogenic |
| | 7:5986933:A:AT | c.1831dupA | p.Ile611fs | 1 | Pathogenic |
| | 7:6003793:C:A | c.251-1G>T | | 1 | Likely pathogenic |
| *APOB* | 2:21006288:C:T | c.10580G>A | p.Arg3527Gln | 1 | Pathogenic |
| | 2:21002392:CAT:C | c.13028_13029delAT | p.Tyr4343fs | 2 | Uncertain |
| | 2:21006128:G:C | c.10740C>G | p.Asn3580Lys | 2 | Uncertain |
| | 2:21005155:TG:T | c.11712delC | p.Asn3904Lysfs | 1 | Pathogenic[b] |
| *LDLR* | 19:11116198:A:G | c.1691A>G | p.Asn564Ser | 1 | Likely pathogenic |
| | 19:11105441:G:A | c.535G>A | p.Glu179Lys | 1 | Likely pathogenic |
| | 19:11120143:C:T | c.1897C>T | p.Arg633Cys | 1 | Pathogenic |
| | 19:11123263:C:T | c.2230C>T | p.Arg744Ter | 1 | Pathogenic |
| | 19:11105567:G:A | c.661G>A | p.Asp221Asn | 1 | Pathogenic |
| | 19:11116153:G:A | c.1646G>A | p.Gly549Asp | 1 | Pathogenic |
| | 19:11113608:G:A | c.1432G>A | p.Gly478Arg | 1 | Likely pathogenic |
| | 19:11113343:G:A | c.1252G>A | p.Glu418Lys | 1 | Uncertain |
| | 19:11120106:G:T | c.1860G>T | p.Trp620Cys | 1 | Uncertain |
| | 19:11120188:T:G | c.1942T>G | p.Ser648Ala | 2 | Uncertain |

| | | | | | |
|---|---|---|---|---|---|
| | 19:11131285:A:G | c.2552A>G | p.Gln851Arg | 1 | Uncertain |
| | 19:11105507:G:A | c.601G>A | p.Glu201Lys | 1 | Uncertain |
| | 19:11105572:C:T | c.666C>T | p.D222= | 1 | Uncertain |
| *TTR* | 18:31598655:G:A | c.424G>A | p.Val142Ile | 31 | Pathogenic |
| | 18:31592974:G:A | c.148G>A | p.Val50Met | 1 | Pathogenic |

[a]Only Pathogenic and Likely Pathogenic variants associated with conditions included in the pilot genomic screening program were disclosed to consenting participants.
[b]This *APOB* variant is associated with hypobetalipoproteinemia.
cDNA and protein position provided for NM_007294.3 (*BRCA1*), NM_000059.3 (*BRCA2*), NM_000249.3 (*MLH1*), NM_000179.2 (*MSH6*), NM_000535.5 (*PMS2*), NM_000384.2 (*APOB*), NM_000527.4 (*LDLR*), and NM_000371.3 (*TTR*); Human reference genome build 38 (GRCh38).

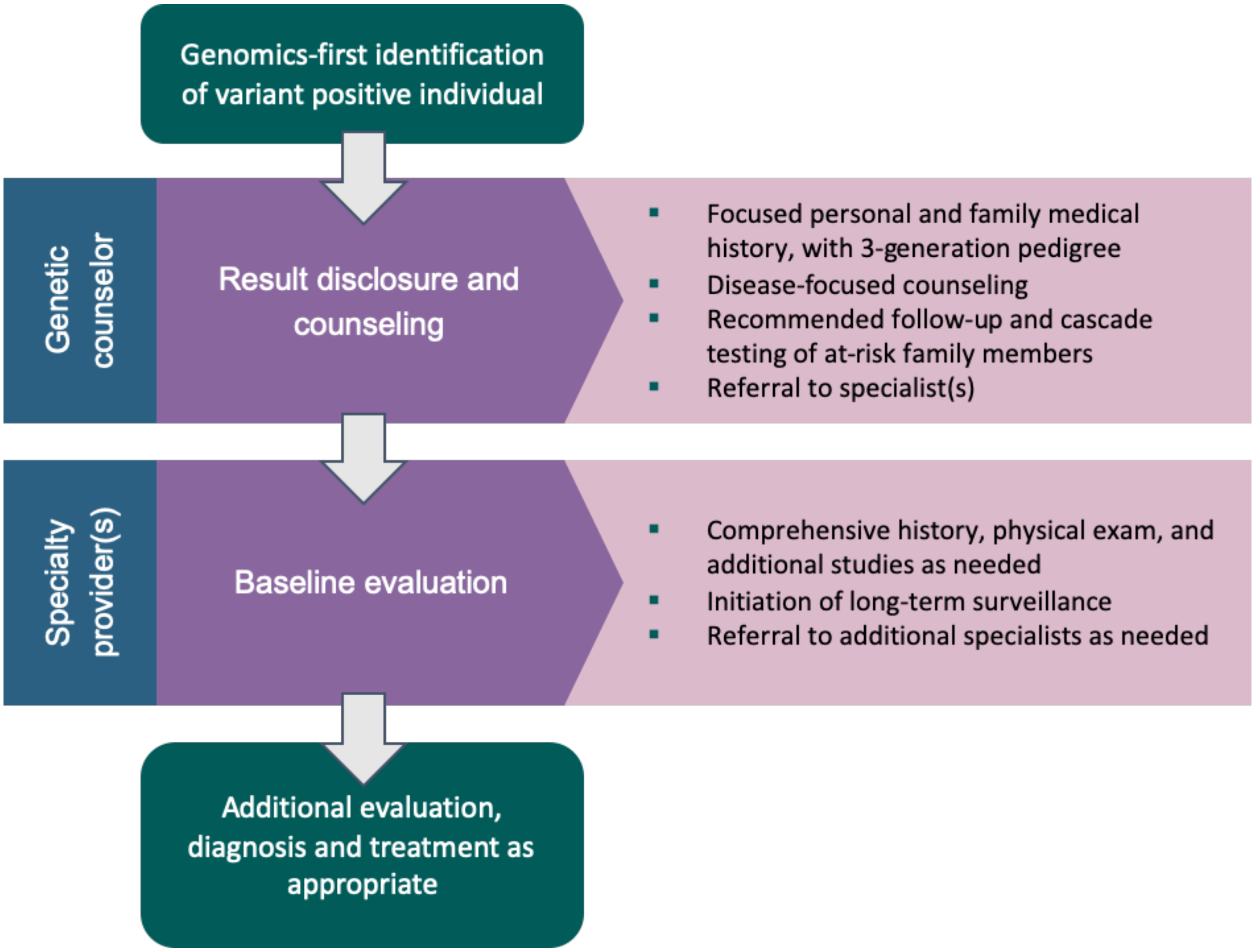


**Figure S1. A model for returning genomic results to biobank research participants without prior knowledge of genomic risk.** After genomics-first identification of variant positive individuals, they receive genetic counseling and are then referred to specialty provider(s) for further evaluation and initiation of risk management.

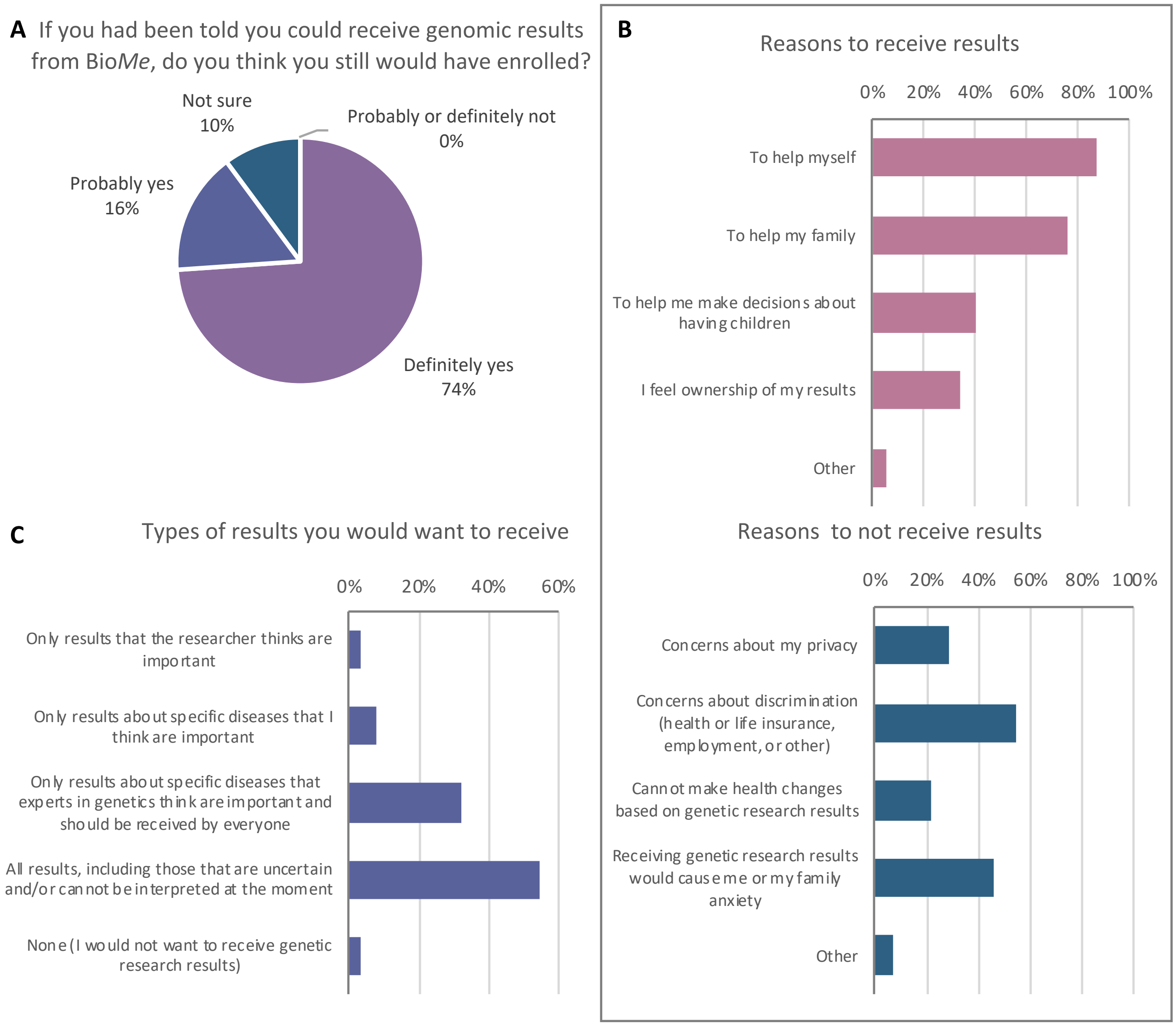


**Supplementary Figure 2. Pre-pilot survey to understand Bio*Me* participants' (N = 72) preferences regarding the hypothetical return of results. A)** Decision to enroll into biobank research if the program returned genomic results. **B)** Selected reasons for receiving or not receiving genomic results. **C)** Selected categories of genomic results participants would want to receive.
